# UV-A and UV-B Can Neutralize SARS-CoV-2 Infectivity

**DOI:** 10.1101/2021.05.28.21257989

**Authors:** Mara Biasin, Sergio Strizzi, Andrea Bianco, Alberto Macchi, Olga Utyro, Giovanni Pareschi, Alessia Loffreda, Adalberto Cavalleri, Manuela Lualdi, Daria Trabattoni, Carlo Tacchetti, Davide Mazza, Mario Clerici

## Abstract

We performed an in-depth analysis of the virucidal effect of discrete wavelengths: UV-C (278 nm), UV-B (308 nm), UV-A (366 nm) and violet (405 nm) on SARS-CoV-2. By using a highly infectious titer of SARS-CoV-2 we observed that the violet light-dose resulting in a 2-log viral inactivation is only 10^−4^ times less efficient than UV-C light. Moreover, by qPCR and fluorescence in situ hybridization (FISH) approach we verified that the viral titer typically found in the sputum of COVID-19 patients can be completely inactivated by the long UV-wavelengths corresponding to UV-A and UV-B solar irradiation. The comparison of the UV action spectrum on SARS-CoV-2 to previous results obtained on other pathogens suggests that RNA viruses might be particularly sensitive to long UV wavelengths. Our data extend previous results showing that SARS-CoV-2 is highly susceptible to UV light and offer an explanation to the reduced incidence of SARS-CoV-2 infection seen in the summer season.

**SYNOPSIS:** UV-A, UV-B and violet wavelengths kill SARS-CoV-2, supporting the sterilizing effects of the solar pump on human pathogens and the explanation of the seasonality of the COVID-19 pandemic.

## INTRODUCTION

At the end of 2019 a new coronavirus, SARS-CoV-2, was firstly described in Wuhan, China, as being responsible for pneumonia within the scenario of a new disease: COVID-19. COVID-19 rapidly became a worldwide pandemic which is still raging and has been responsible for dramatic and unforeseeable health, social and economic consequences^1^. The development of multiple SARS-CoV-2 vaccines made it possible to predict that the pandemics will eventually be defeated^2–4^, but the implementation of disinfection and prevention strategies is still of the foremost importance to curb the spread of new infections^5–7^.

The germicidal effect exerted by UV light on bacteria and viruses has been widely documented since the end of 1800^8^. UV-C light produced from low pressure mercury tubes is commonly employed in the disinfection of wastewaters and closed environments as well as of blood products, and represents a well-established, economical and safe technology^9–13^. SARS-CoV-2 was recently shown to be highly sensitive to UV-C light^14–16^, although discrepancies in the results indicated that all the variables involved in the experimental setting have to been taken into account to obtain reliable and replicable data^14^. These results are of paramount importance for the development of air and surface disinfecting devices, but do not explain the particular seasonal epidemiology of SARS-CoV-2 infection which follows a bimodal temporal pattern, peaking in winter and being greatly reduced in summer. Thus, the idea that viral epidemiology could be modulated by the intensity of the solar pump is rebutted by the observation that the UV-C light emitted by the Sun, the most important UV light source, is filtered and blocked by the ozone layer in the stratosphere. UV-A and UV-B radiations, on the other hand, reach the earth surface with different intensities, which depend on the season, the latitude, and the weather conditions. The effect of solar UV-A and UV-B radiation on microorganisms^17,18^ and on the seasonal behavior of infectious diseases has been extensively discussed^19,20^, and recent predictive models suggest that SARS-CoV-2 infection is indeed solar-sensitive^21–23^. Solid, convincing and reproducible experimental data on the possible virucidal effects of UV-A and UV-B and their correlation with SARS-CoV-2 epidemiology are nevertheless missing.

Here, we used the TCID_50_ approach to quantify SARS-CoV-2 inhibition upon irradiation, using wavelengths spanning form UV-C to violet light and we build the corresponding UV-action spectrum. Next, we used PCR and FISH measurements to identify the UV-A and UV-B doses that completely inhibit the viral concentration comparable to the one present in the sputum of SARS-CoV-2-infected patients. Finally, we compared the obtained UV action spectrum on SARS-CoV-2, with those reported in the literature on other viruses and bacteria.

Our data define the wavelengths and the doses of UV radiation that result in SARS-CoV-2 inactivation and show that SARS-CoV-2 infectivity can be completely blocked by UV-A and UV-B. These results offer an explanation to the seasonality of such infection, and provide fundamental information to build sterilization devices that avoid the toxicity of UV-C-based methods and that could be potentially effective in the inhibition of multiple RNA viruses.

## MATERIALS AND METHODS

### Analyses of SARS-CoV-2 replication following UV-exposure

2 × 10^4^ VeroE6 cells were cultured in DMEM (ECB20722L, Euroclone, Milan, Italy) with 2 % FBS medium, 100 U/ml penicillin, and 100 μg/ml streptomycin, in a 96-well plate for 24 hours before infection. To determine SARS-CoV-2 susceptibility to UV-irradiation, a viral stock (Virus Human 2019-nCoV strain 2019-nCoV/Italy-INMI1, Rome, Italy) at a concentration of 6 × 10^6^ TCID_50_/mL in DMEM (ECB20722L, Euroclone, Milan, Italy) was placed under the LED lamp and irradiated with 3 different doses (D1, D2, D3) for each tested UV-wavelength (see Table 1). For each UV-wavelength viral titers were determined by TCID_50_ endpoint dilution assay. Briefly: serial ten-fold dilutions, from 10^6^ to 10^−4^ TCDI_50_/mL (50ul), were plated onto 96-well plates, incubated at 37°C in 5% CO_2_ and checked daily to monitor the UV-induced cytopathic effect. Seventy-two hours post infection (hpi) viral titer was determined as previously described^24^.

**Table 1.** Features of the illumination conditions. Transmittance through a 1mm thick DMEM layer at the selected wavelengths; the three doses provide to the virus stock for the same wavelengths.

| LED | Peak Wavelength (nm) | DMEM |  | Dose (mJ/cm<sup>2</sup>) |  |  |
| --- | --- | --- | --- | --- | --- | --- |
|  |  | Transmittance |  | D1 | D2 | D3 |
|  |  | Min | Ave |  |  |  |
| UV-C | 278 | 0.647 | 0.811 | 2 | 4 | 12 |
| UV-B | 308 | 0.867 | 0.932 | 100 | 200 | 600 |
| UV-A | 366 | 0.901 | 0.950 | 2000 | 4000 | 12000 |
| Violet | 405 | 0.897 | 0.948 | 4000 | 8000 | 24000 |

The same UV-wavelengths were used to irradiate a SARS-CoV-2 viral stock at a concentration of 1.5 × 10^3^ TCID_50_/mL. VeroE6 cell cultures were incubated with the virus inoculum in quadruplicate for one hour at 37°C and 5% CO_2_ and viral replication was assessed by qPCR, as previously described^14^, at 24, 48 and 72 hpi. SARS-CoV-2 that was not exposed to UV light was used as control.

### Fluorescence in-situ hybridization (FISH)

It was performed using previously described 96 smiFISH probes^25^ that target the whole (positive-strand) genome of the virus. According to the smiFISH protocol^26^, a FLAP sequence is appended at the 3’ of each primary probe. Prehybridization with a complementary Cy3-labelled Cy3 probe was performed as using a PCR machine, as described^25^.

A SARS-CoV-2 viral stock (1.5 × 10^3^ TCID_50_/mL) was exposed to the UV-doses reported in Table 1 and an *in vitro* infection assay was performed on 1×10^5^ Vero E6 cells grown on a 13mm glass coverslip (0.17mm thickness) as described in the previous section. Twenty-four hpi the supernatant was collected to quantify viral replication by qPCR, while cells were fixed by 4% PFA for 10 minutes, washed twice with PBS, and then stored until labeling in 70% ethanol at −20°C (minimum ethanol incubation: overnight). The day of the labeling, the coverslips were brought to room temperature, washed twice with 10% SSC-20× in RNase-free water (Buffer I) for 5 minutes, followed by a wash in 10% SSC-20× and 20% formamide in nuclease-free water (Buffer II). Cells were incubated overnight with the hybridized FLAP-duplex in a humidified chamber at 37°C. The probes were diluted 1:100 in the hybridization buffer (10% (w/v) of dextran sulfate, 10% of SSC-20× buffer and 20% formamide in Rnase-free water). Following hybridization, cells were washed twice in Buffer II in the dark for 30 min at 37°C, then washed in PBS for 10 min and stained with 1μg/ml Hoechst 33342 in PBS. The coverslips were then mounted on glass slides using Vectashield (Vector Laboratories, Peterborough, UK).

Images were acquired on a custom-built epifluorescence microscope, equipped with a led source for illumination (Excelitas Xcite XLED1, Qioptiq, Rhyl, UK), an Olympus LUCPlanFLN 20x/0.45NA objective (Olympus Life Science Segrate, IT) and a Hamamatsu Orca Fusion sCMOS detector (Hamamtsu Photonics Italia S.R.L, Arese, Italy), resulting in a pixel size equal to 324.6nm. By using a motorized stage (Mad City Labs GmbH, Schaffhauserstrasse, CH), mosaic of 6×6 fields of views were collected and then stitched using the Image Stitching ImageJ plugin^27^. The resulting large fields of view (approx. 3mm of side) were then scaled by a factor 0.25 in FiJi^28^ and then imported in Matlab for image analysis using a custom-written routine. The code segments the nuclei in the Hoechst channel – discarding objects smaller than 40 pixels or larger than 400 pixels – and then computes the average intensity in the FISH probes channel, in a 3-pixel wide ring surrounding each individual nucleus. The distribution of the single-cell FISH signal intensity is then plot as a histogram. The distribution of the single-cell FISH signal intensity is then plot as a histogram and the fraction of positive cells is calculated by counting those cells with a FISH signal intensity 2^3^ times higher than background intensity.

### LED illumination conditions

To irradiate the virus we placed the above mentioned virus titers in 6-well plates, which were then irradiated using LEDs with the spectral features reported in Figure 1A. While these sources are not monochromatic as in the case of low-pressure mercury lamps, they show a bandwidth of about 10 nm, similar to the sources used to generate the action spectrum on other viruses^29^. The sources were calibrated using an Ocean Optics HR2000+ spectrometer (Ocean Optics Inc., Dunedin, USA). The spectrometer was calibrated against a reference deuterium–halogen source (Ocean Optics Inc. Winter Park, Winter Park, Florida) and in compliance with the National Institute of Standards and Technology (NIST) practices recommended in NIST Handbook 150-2E, Technical guide for Optical Radiation Measurements. The last calibration was performed in March 2019. During irradiation the virus is suspended in a 1mm thick layer of DMEM medium: in the UV-range, medium absorption is non-negligible and needs to be accounted to calculate the real dose delivered to the virus^14^. It is important to note that the virus itself does not contribute significantly to the source attenuation instead, as its concentration is too low to have a significant effect (see results). To compute the fraction of UV light that is absorbed by the medium we used the absorption spectrum of DMEM reported in Figure 1B and calculated the transmittance trend as function of the liquid thickness (Figure 1C). The resulting fraction of UV light delivered on average in the well (average) and at the bottom of the well (minimum) as function of the wavelength is reported in Table 1.

**Figure 1.**
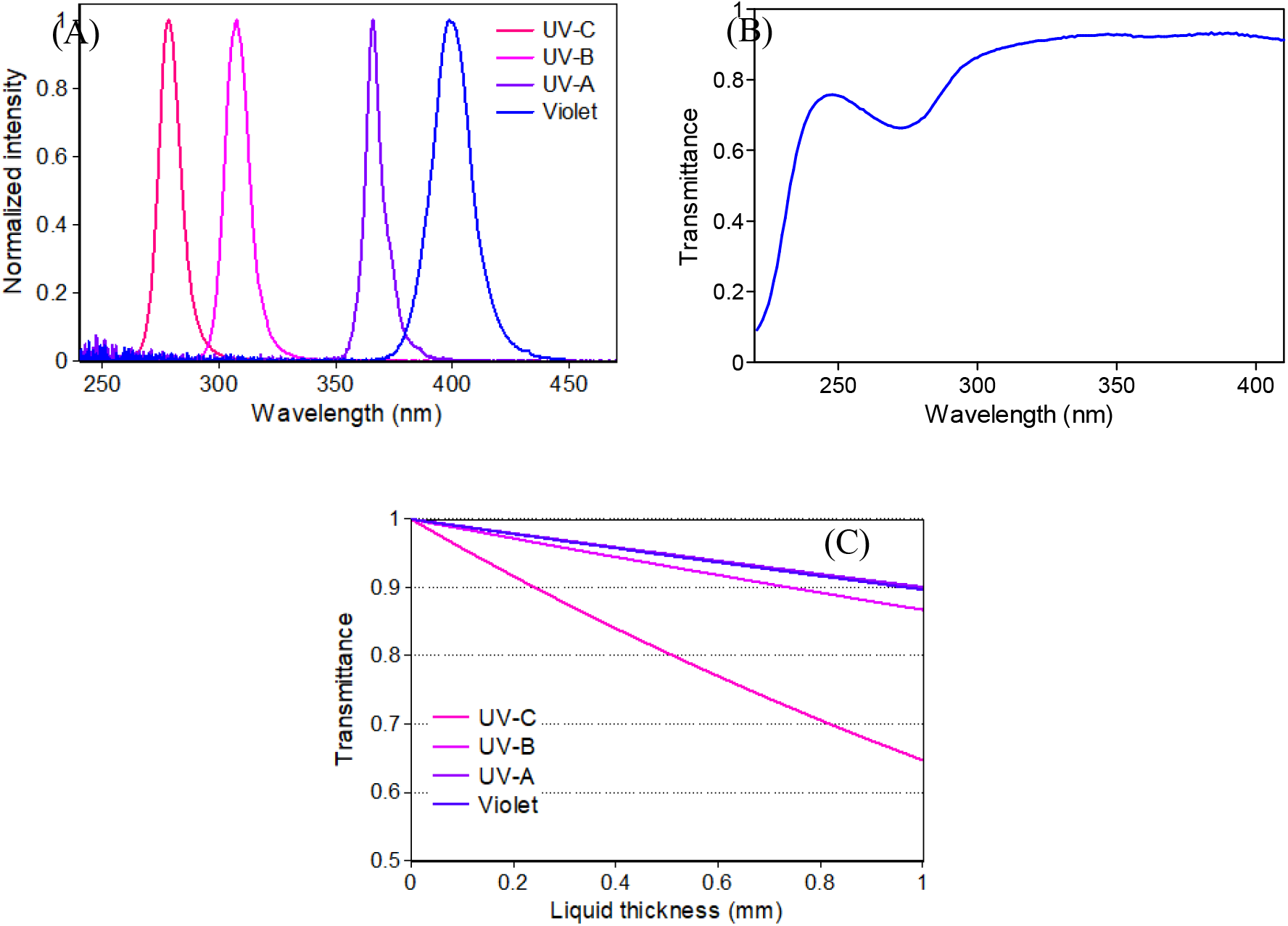
Features of the illuminated medium. A) Normalized emission spectra of the LEDs used in the experiments. B) Transmission spectrum of the DMEM used in the experiment (1 mm thick). C) Fraction of transmitted UV light as function of the liquid thickness.

**Figure 2.**
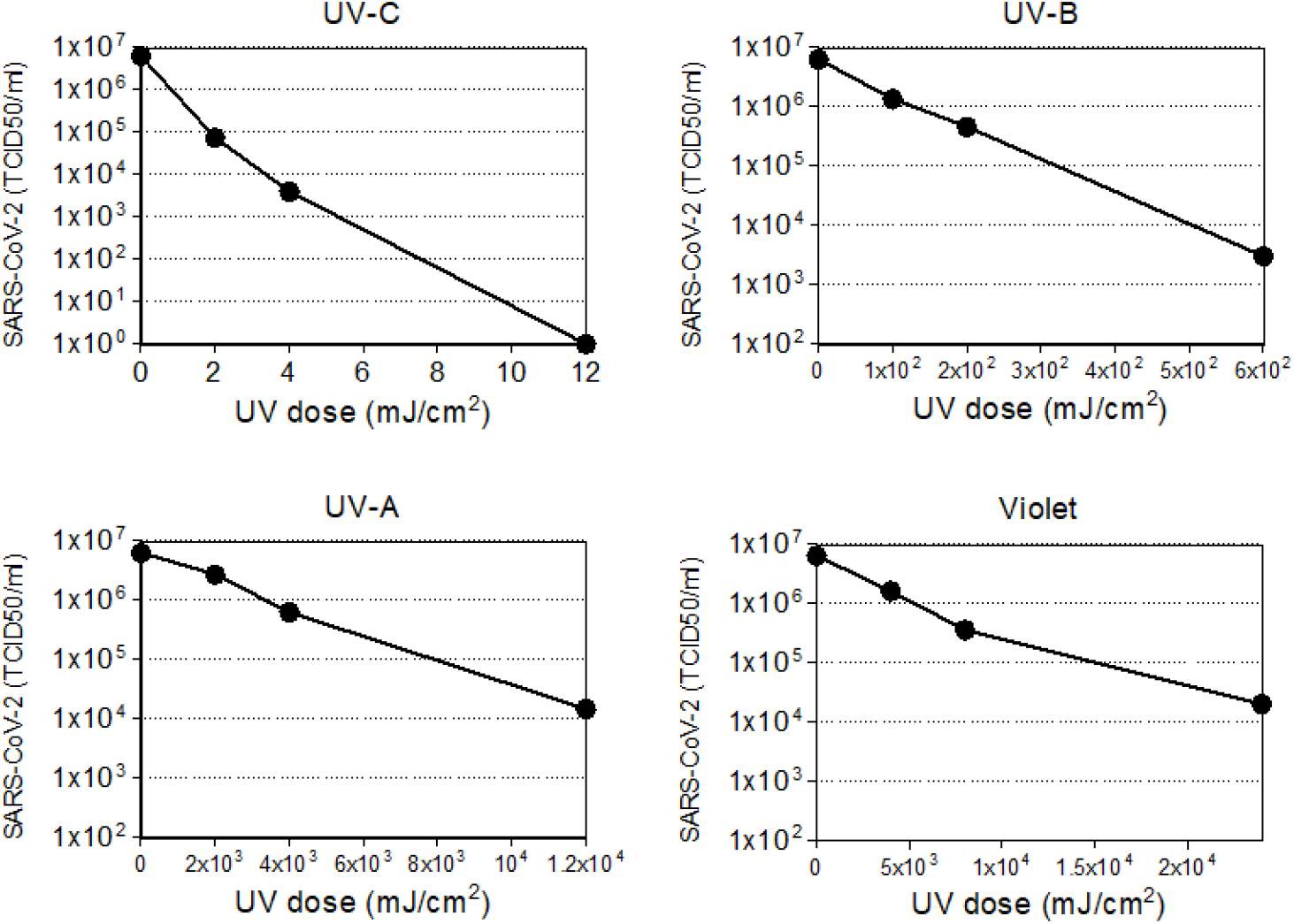
Virus titer trend as function of the light doses. The trend is reported for the four different wavelengths tested (UV-C,-B,-A stand for 278, 308, and 366 nm respectively, violet for 405 nm).

As expected from the transmittance spectrum, for UV-A and UV-B light the minimum and average values were comparable (approx. 90-95%), indicating that all the virions suspended in the solution do receive the same UV dose. At 278 nm (UV-C), the discrepancy between the minimum and average value became more evident. The consequence is that the viruses at the bottom of the well receive 20% less UV photons than the average.

According to the spectral and irradiance calibrations and the DMEM transmittances, the exposure times were calculated to provide the doses reported in Table 1.

### Statistical Analyses

In the bar plots in Figure 3, the mean ± standard error of the mean (SEM) are indicated. Data were analyzed using two-way ANOVA test by GRAPHPAD PRISM version 5 (Graphpad software, La Jolla, Ca, USA), and p-values of 0.05 or less were considered to be significant.

**Figure 3.**
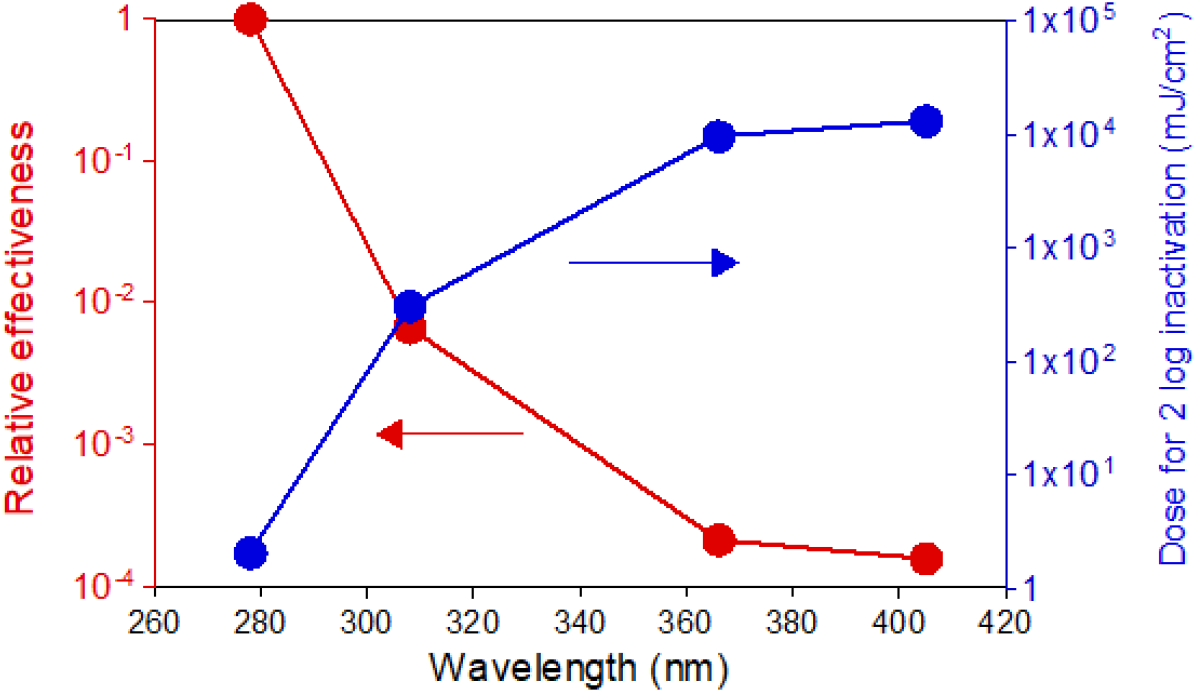
SARS-CoV-2 action spectrum according to the TCID50 measurements. The plot reports the spectrum normalized at 1 for the UV-C (red curve) and the dose required for a 2-Log inactivation (blue curve).

## RESULTS AND DISCUSSION

We first applied the TCID_50_ approach to measure how effective UV light of different wavelengths is in inactivating the virus. To achieve this result, we infected cells with an initial viral concentration (6 × 10^6^ TCID_50_/mL), which is significantly higher than that observed in infected patients. Next, we performed experiments on lower initial viral loads, comparable to the ones present in the sputum of COVID-19 samples^30^. Here, we used qPCR to evaluate the capability of the irradiated virus to replicate and FISH to quantify the ability of the different UV doses to impair SARS-CoV-2 infectivity at the single-cell level.

### Wavelength-dependent inhibition of SARS-CoV-2 infectivity

Results reported in Figure 2 describe the trend of the viral titer as function of the dose for the four wavelengths considered.

For all the analyzed wavelengths, we observed a dose-response relationship between the administered UV-dose and viral titer, indicating that virus is susceptible to different extents to a wide range of UV wavelengths. Notably, the decreasing trend was observed to be exponential, as predicted for such kind of mechanism^31^. Importantly, while a few mJ/cm^2^ of UV-C light are sufficient to cause a several order of magnitude drop in the effective viral concentration, 10x to 1000x higher doses were needed to observe a similar viral load reduction following UV-B and UV-A irradiation.

These data confirm earlier findings indicating that UV-C is more effective in virus inactivation compared to both UV-B and UV-A, and reinforce the possibility of using UV-C irradiation as an efficient method for a fast inactivation of SARS-CoV-2^14–16^. Further, the data indicate that UV-B and UV-A also have virucidal potential, although at higher doses than UV-C.

### Action spectrum

To better characterize these results, we built the action spectrum at a 2-Log inactivation of SARS-CoV-2. We extrapolated the data fitting the effective concentration curves of Figure 2 with a simple single exponential trend, normalized the action spectrum by setting at 1 the inhibition efficiency at 278 nm and calculated the relative effectiveness at the other wavelengths (Table 2 and Figure 3).

**Table 2.** Extrapolated doses resulting in a 2-log reduction. The data are derived according to the TCID50 measurements and the relative efficiency as function of the wavelength.

| Wavelength<br>(nm) | 2-Log dose<br>(mJ/cm<sup>2</sup>) | Relative<br>effectiveness |
| --- | --- | --- |
| 278 | 2.07 | 1 |
| 308 | 309.07 | 4.2x10<sup>-3</sup> |
| 366 | 9719.65 | 2.0x10<sup>-4</sup> |
| 405 | 13224.56 | 1.1x10<sup>-4</sup> |

The data and the plot recapitulate the results obtained by TCID_50_, namely that the measured relative effectiveness of UV irradiation on SARS-CoV-2 reaches a plateau at longer wavelengths, with UV-C being 10^4^ times more effective than violet light.

Such a substantial difference is expected given that the absorption spectrum of both proteins and nucleic acids peak in the UV-C region. Nevertheless, the result that higher doses of UV-A and violet light can still have a virucidal effect on SARS-CoV-2 is noteworthy, as it supports the idea of a sterilizing effect of solar radiation on the virus.

### UV-dependent virus inhibition of SARS-CoV-2 infectious titers equivalent to those present in COVID-19 patients

To examine the efficacy of UV-inactivation in a real-world scenario, we used the UV-doses reported in Table 1 on a SARS-CoV-2 viral concentration equivalent to the one found in the sputum of SARS-CoV-2 infected patients (1.5×10^3^ TCID_50_/ml)^30^. QPCR was employed to quantify viral replication over time (Figure 4).

**Figure 4.**
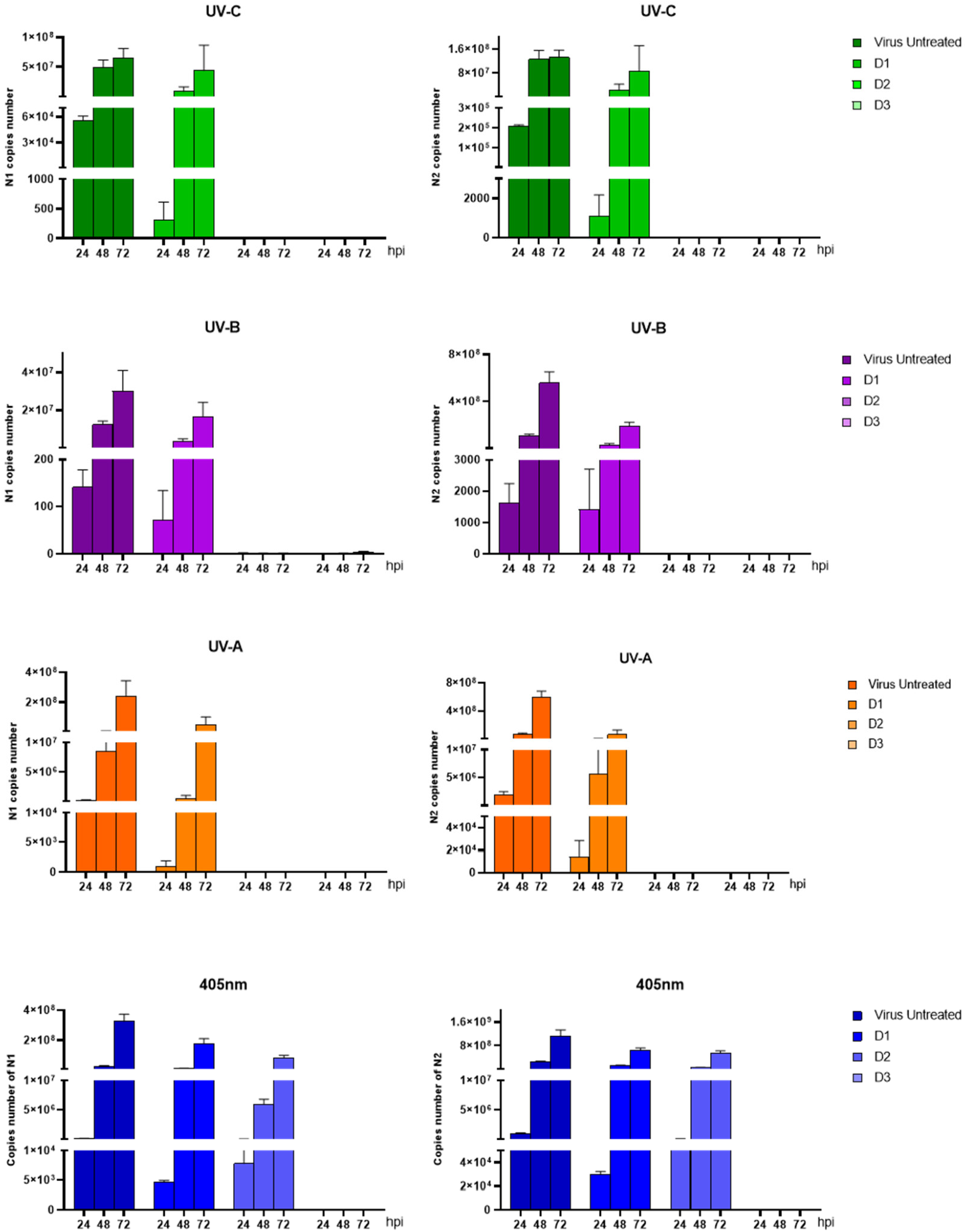
Viral replication of UV-irradiated SARS-CoV-2 (1,5×106 TCID50) in the supernatant of in vitro infected VeroE6 cells. Vero E6 cells were infected with SARS-CoV-2 irradiated with different doses (D1, D2, D3) of UV-A, -B, -C and violet light. Culture supernatants were harvested at the indicated times (24, 48 and 72 hpi) and virus titers were measured by absolute copy number quantification (Real-Time PCR). All cell culture conditions were seeded in quadruplicate. Mean values ±SEM are shown.

Results showed that all the wavelengths analyzed are capable to completely inhibit the amplification of the viral genome though, as expected, with different efficacy. In particular, a marked reduction of viral copy number was observed at 24 hour post infection (hpi) with a low dose of UV-C (278 nm) equal to 4 mJ/cm^2^. Even more important, the viral copy number did not increase over time (72 hpi), endorsing the complete inhibition of the virus. This is in line with the results reported with the 254 nm light^14^ and confirms the high susceptibility of the virus to UV-C photons. Notably, and most interestingly, results indicated that a 200 mJ/cm^2^ UV-B dose, a 4000 mJ/cm^2^ UV-A dose and a 24000 mJ/cm^2^ violet dose were also sufficient to completely inactivate the virus.

All the results were further confirmed by 2-ANOVA statistical analyses performed on viral replication at extracellular level for UV-C (4 mJ/cm^2^ vs. untreated: N1, p = 0,0243; N2: p = 0,0119; 12 mJ/cm^2^ vs. untreated: N1, p = 0,0243; N2: p = 0,0119), UV-B (200 mJ/cm^2^ vs. untreated: N1, p = 0,0009; N2: p = <0,0001; 600 mJ/cm^2^ vs. untreated: N1, p = 0,0007; N2: p = <0,0001), UV-A (4000 mJ/cm^2^ vs. untreated: N1, p = 0,0283; N2: p = <0,0001; 12000 mJ/cm^2^ vs. untreated: N1, p = 0,0233; N2: p = <0,0001) and 405nm (4000 mJ/cm^2^ vs. untreated: N1, p = 0,0033; N2: p = 0,0056; 8000 mJ/cm^2^ vs. untreated: N1, p = <0,0001; N2: p = 0,0002; 24000 mJ/cm^2^ vs. untreated: N1, p = <0,0001; N2: p = <0,0001) (Figure 4).

By comparing these results with those based on TCID_50_, we clearly observed that viral concentration is a key variable in the experimental setting. For example, a 200 mJ/cm^2^ UV-B dose is sufficient to totally inhibit viral replication at a viral concentration comparable to the one in the sputum of a COVID-19 patient, but results in just a 1-log reduction of the higher viral titer. This feature is crucial to properly compare different studies with different experimental conditions, in particular if these data should be used to develop models of sun irradiation on virus dispersed as aerosol in the atmosphere. It is important to note that the viral concentration in such case is expected to be much lower than those used in our experimental tests.

### FISH experiments

In order to directly visualize the capability of UV to interfere with the SARS-CoV-2 infection of host cells, we next performed an *in vitro* infection assay on VeroE6 cells following incubation with a 1.5×10^3^ TCID_50_/ml virus titer that had or had not been irradiated with different UV wavelengths. FISH probes targeting the positive strand of the viral RNA (vRNA) were used in the experiments. Time-course analyses using non-irradiated virus showed that the FISH signal progressively increases in single cells over time due to the replication of the viral genome, and shifts from a punctate staining (representative of few vRNA copies/cell), which is observed at early time-points, to an intense cytosolic signal at 24hpi (Figure 5A). Such robust activation at late time points allows to detect SARS-CoV-2 positive cells with low magnification objectives, thereby enabling the analysis of thousands of cells per condition (Figure 5B-C).

**Figure 5.**
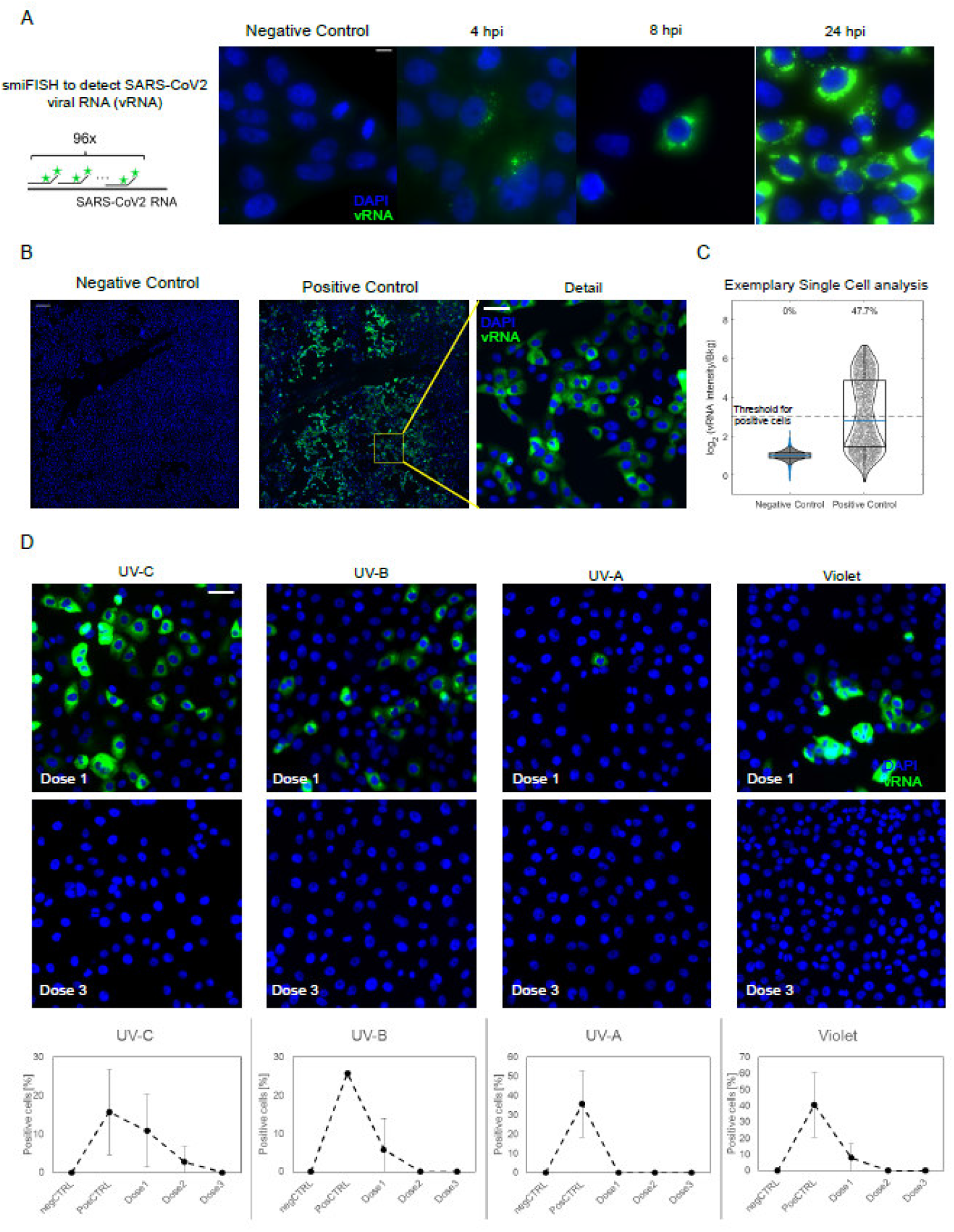
Fluorescence In-situ hybridization (FISH) of the SARS-CoV-2 viral RNA (vRNA) in VeroE6 cells. (A) Scheme of the hybridization of the FISH probes (left) and timecourse of SARS-CoV-2 infection at different timepoints post infection, as collected with an high magnification/Numerical aperture (NA) objective (60x 1.49NA). Scale bar 10❍m. (B) The bright signal of the smFISH probes at 24hpi allow to use a low NA objective (20x, 0.45 NA) to reliably identify infected cells among thousands of cells, by collecting 6×6 mosaics (total field of view approx. 3mm, scale bar in the ‘detail’ image 50 μm).(c) Positive cells are identified as those with a 8-fold higher intensity I than the background n the FISH channel. (D) Exemplary details of 6×6 mosaics upon irradiation with different wavelength and doses of UV/Violet light. UV doses are as indicated in Table 1. Quantification is provided as mean +/-standard deviation of the fraction of vRNA positive cells on at least two 6×6 mosaics. Scale bar: 10 μm.

Cell-by-cell distribution of the FISH signal displayed a bi-modal distribution, with 20-40% of cells being clearly infected at 24 hours post infection when non-irradiated virus was seeded on cells (Figure 5C). In contrast with these data, all the analyzed UV-wavelength reduced the fraction of positive cells, as probed by smFISH, resulting in a complete inhibition of the virus when SARS-CoV-2 was irradiated with wavelength specific critical doses (4mJ/cm^2^ for UV-C, 100mJ/cm^2^ for UV-B, 4000 mJ/cm^2^ for UV-A and 24000 mJ/cm^2^ for violet light) (Figure 5D). Notably at lower UV doses some cells with high levels of vRNA could be detected, suggesting that, rather than inhibiting the capability of the viral genome to replicate, UV-irradiation might affect either virus entry in the cells or the assembly of functional viral particles following the replication of the viral genome.

### Action-spectrum comparison of the UV-susceptibility of different microorganisms

We finally compared the results obtained in the TCID_50_ assay with those reported in the literature for other viruses and bacteria by building the UV action spectra for each of the different pathogens (Figure 6). Data were normalized to the UV-C inactivation dose (for SARS-CoV-2 it has been assumed the same level of inactivation at 253 and 278 nm).

**Figure 6.**
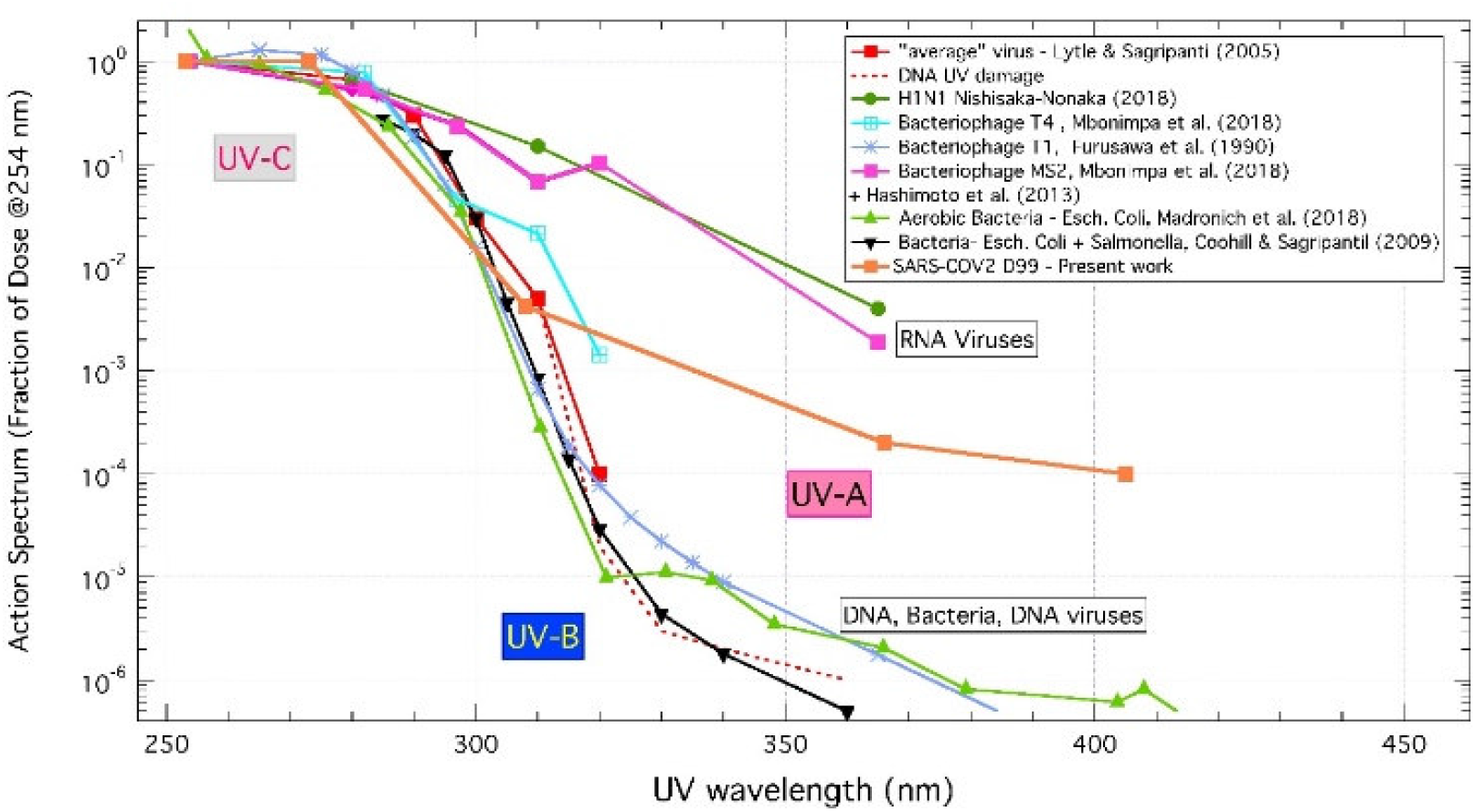
Action spectra from UV-C to UV-A for different microorganisms (bacteria and viruses) and DNA. They’re inferred by previous works with the results achieved in the present work for SARS-CoV-2 normalized to the UV-C inactivation dose (see text for details). The data refers to inactivation dose levels (between 1-Log and 2-Log).

The viruses we included in the analysis are influenza virus H1N1 ^29^ and the Bacteriophages MS2^32,33^, T1^34^ and T4^33^. The “average” virus UV action spectrum inferred by Lytle and Sagripanti^19^ and Horton et al.^35^ and combining data achieved with different viruses (see also ref^35^) are shown as well. For bacteria, results for Escherichia Coli alone or combined with Salmonella^36,37^ are presented. DNA-damage action spectra are also displayed^38^.

It should be noted that whereas for both bacteria and DNA viruses the inactivation observed in the UV-A is 10^−5^-10^−6^ times lower than that observed in the UV-C region, for RNA viruses the inactivation power is only marginally lower in the UV-A than in the UV-C region (just about 100-1000 times lower at 366 nm). Taking into account that the Solar illumination in the UV-A region is much larger than in the UV-B (95% of UV-A and 5% of UV-B), these results might justify the acknowledged seasonal behavior of the outbreaks of airborne viruses, including corona^39–41^ and influenza viruses^42,43^. In this respect, we would like to stress how it is crucial to have the specific data of the target microorganism to develop reliable solar inactivation and/or seasonal models and how Lytle and Sagripanti action spectrum cannot be considered as a reference trend.

## CONCLUSIONS

The ability of UV light to inactivate SARS-CoV-2 replication was studied as function of the irradiation wavelengths. Results showed that irradiation of a viral stock with a high infectious titer irradiated with LEDs at 278, 308, 366 and 405 nm resulted in a significant decrease in the fraction of active virus proportional to the light dose for all the wavelengths. Considering a 2-Log decrease and fixing to 1 the efficiency at 278 nm, it was possible to build the action spectrum, which is characterized by an effectiveness ratio of 10^−4^ at 405 nm, indicating that even violet light has a non-negligible efficiency in inactivating SARS-CoV-2. While such a relatively flat action spectrum might appear surprising, our comparison of multiple UV-action spectra on diverse microbes revealed that RNA viruses, including SARS-CoV-2, might be more susceptible than other pathogens to long wavelength UV-light.

These observations were confirmed by experiments performed on a virus titer equivalent to that present in COVID-19 patients. Thus, in these experimental conditions it was possible to completely inhibit virus replication with only 4 mJ/cm^2^ at 278, 200 mJ/cm^2^ at 308 nm, 4000 mJ/cm^2^ at 366 nm and 24000 mJ/cm^2^ at 405 nm. FISH analysis confirmed that UV-C,UV-B, UV-A, and violet light irradiation can inhibit SARS-CoV-2 infection of target cells These results are possibly extremely relevant as they analyze the effects of different UV wavelengths on SARS-CoV-2 titers that relate to the real life scenario of the current pandemic. In particular, data herein indicate that solar irradiation reaching the surface of the Earth could completely inactivate a titer of isolated SARS-CoV-2 similar to the one found in the sputum of COVID-19 patients within minutes. In conclusion, for the first time, we have demonstrated that UV irradiation is effective in SARS-CoV-2 inhibition at multiple wavelengths including UV-A and violet light. These results support a role for solar irradiation in viral disinfection of external surfaces and might contribute in explaining the seasonality of this virus.

## Data Availability

All data are published in the manuscript

## AUTHOR INFORMATION

### Author Contributions

The manuscript was written through contributions of all authors. All authors have given approval to the final version of the manuscript.

## ACKNOWLEDGMENT

We are grateful to De Sisti Lighting for having provided a custom designed multi-LED UV lamp used in the experiments. We are grateful to Dr. F Mueller and Dr. C. Zimmer (Institut Pasteaur, Paris) for kindly providing the smiFISH probes. We acknowledge F. Nicastro, J. R. Brucato, P. Tozzi, I. Ermolli, G. Sironi, E. Antonello from INAF for many useful discussions.

